# Behavioural determinants of testing behaviour during a hypothetical avian influenza outbreak: an interview study

**DOI:** 10.64898/2026.03.17.26348610

**Authors:** Rosa C. van Hoorn, Laurens C. van Gestel, Diana S. Griffioen, Mariska W.F. Petrignani, Coco Kersten, Margriet Müskens, Laura Vols, Hanneke Borgdorff, Irene M. van der Meer, Marieke A. Adriaanse, Adriënne S. van der Schoor

## Abstract

**Background:** Avian Influenza (AI) is a potential pandemic threat, specifically when human-to-human transmission occurs. For outbreak management testing is essential. Current knowledge on testing behaviour is mostly derived from other infectious diseases such as COVID-19. It is necessary to identify determinants of testing behaviour for AI in an early phase. Therefore, this interview study aims to identify a wide range of behavioural determinants of testing during a hypothetical human-to-human transmissible AI outbreak.

**Methods:** Semi-structured in-depth interviews, based on the Theoretical Domains Framework, were carried out between May 2024 and February 2025. Participants were included through purposive and convenience sampling. During the interviews an animation was shown illustrating a hypothetical AI outbreak. Verbatim transcripts were thematically analysed.

**Results:** We included seventeen participants (median age 44, range 20-81; 71% women) with diverse backgrounds in terms of age, gender, educational level and country of birth. We found that having the freedom to decide to test would make testing more acceptable, whereas a decreased sense of autonomy would discourage testing. Most themes included individual rather than population-level benefits as drivers of testing behaviour. These included protecting loved ones, one’s own health and gaining psychological reassurance. External conditions like being unable to go to work or an event would generally encourage testing behaviour. Lower trust in governmental authorities could hamper testing behaviour. Previous experiences from the COVID-19 pandemic shaped participants’ answers about AI testing behaviour.

**Conclusion:** Key considerations include balancing people’s need for autonomy with the external measures imposed by employers or the government, rebuilding trust in institutions and acknowledging how prior experiences with testing may shape testing behaviour in future AI outbreaks. Further research is needed to determine how these findings can be translated into effective communication and how trust in authorities can be build.

## Background

With the growing world population, interactions between wild animals, livestock and humans occur increasingly often. Combined with large-scale human and animal mobility, infectious diseases can spread rapidly (1, 2). According to the World Health Organisation’s (WHO) updated list of emerging pathogens, influenza A viruses, including avian influenza (AI), are a growing concern for public health. Mainly due to the risk of mutating into strains that can efficiently transmit from human to human (3, 4). Although human-to-human transmission currently is rare, European public health agencies and scientists are concerned that AI viruses could adapt further (4–7). The WHO and European Centre for Disease Control (ECDC) highlight the importance of developing strong preparedness plans that include strategies on public health surveillance and effective prevention and control measures, such as testing and vaccination (3, 7).

Testing is essential for timely detection and diagnosis of AI infections, identification of virus subtypes, and management of outbreaks. Beyond diagnosis, testing supports public health surveillance and is required for implementing effective control measures such as contact tracing and case isolation, to prevent disease complications and to interrupt transmission (8–10). Previous evidence from the COVID-19 pandemic indicates that timely testing, when accompanied by other preventive measures such as isolation and treatment, has a considerable impact on the reduction of transmission and improvement of public health outcomes such as the number of incident cases and the mortality rate in the population (11, 12).

High levels of public participation in testing are necessary to contain and mitigate the spread of a virus. However, securing this engagement remains a well-known challenge (13). During the COVID-19-pandemic in the Netherlands, Public Health Services faced fluctuating test rates over time and differences across populations for professionally administered tests, which complicated outbreak control (14, 15). These fluctuations had several causes, including trust in test results, shifts in testing policy or uncertainties when and where to test (16, 17). Moreover, testing behaviour was influenced by the implications attached to the test result, such as worries about isolation as a consequence of a positive test result (16) or in periods when testing was linked to access to social activities.

Previous research provided valuable insight into testing across populations and determinants that may influence testing behaviour. Important determinants are, among others, perceived usefulness of testing (16, 18–22), knowledge and perceptions on the severity of the disease (21–25), perceived disease susceptibility of people within someone’s social network or environment (16, 19, 21) and alignment with social norms (16, 19). Examples of barriers for testing behaviour include expected test invasiveness (19, 22, 26, 27), difficulties with online appointment systems (19, 26), limited accessibility of testing sites (19, 26, 27) and unclear information on testing (16, 27). In situations that are uncertain, such as pandemics, trust in authorities could also play a role as a lack of trust might lead to a lower willingness to test (19, 21). Altogether, these results reveal several important determinants of testing behaviour identified during previous infectious disease outbreaks, particularly the COVID-19 pandemic. However, as new outbreaks may come with different levels of severity and transmissibility and may occur in different societal contexts (28), the extent to which these determinants apply to future outbreaks remains uncertain.

To adequately prepare for future potential hazards like human-to-human AI, there is a necessity of targeted research to identify determinants of testing behaviour in an early phase of such an outbreak. These insights help improve future testing strategies and shape communication campaigns to stimulate testing, so that testing can effectively be used by public health institutions to mitigate public health risks of an outbreak. Therefore, this qualitative study aimed to identify a wide range of behavioural determinants of testing behaviour, when professionally administered, during a hypothetical AI outbreak transmissible from human to human. Using a behavioural science approach, we aimed to obtain a comprehensive and in-depth set of behavioural determinants of testing behaviour.

## Methods

The methods of this study are reported in accordance with Consolidated criteria for Reporting Qualitative studies (COREQ) reporting guidelines (29) (see supplementary information).

### Study design

A qualitative study involving semi-structured in-depth interviews was conducted to explore behavioural determinants related to testing behaviour. During the interview, an animation video was shown illustrating a hypothetical AI outbreak. Participants were interviewed with a list of predetermined questions (see supplementary information) and a supportive visual aid to explain questions visually.

### Recruitment and participants

Participants were recruited using a combination of purposive and convenience sampling strategies to obtain a heterogenous sample in terms of age, gender, educational level and country of birth. Additionally, variation in testing behaviour for infectious diseases in general was considered during the recruitment process by asking participants about their general willingness to test during infectious disease outbreaks (willing/hesitant/unwilling). Recruitment took place in two Public Health Service regions in the province of South Holland. Participants were approached by health promoters working within local community initiatives, direct recruitment on local markets, announcements in Dutch and English via regional public health social media channels, and via community applications (i.e. apps facilitating communication in neighbourhoods).

### Data collection and reflexivity

Data were gathered through individual semi-structured in-depth interviews to keep consistency across the interviews while also have the flexibility to explore the participants’ perspective. The interview guide was developed based on the TDF, to ensure a comprehensive perspective (30, 31). This framework consists of 14 theory-based domains that represent various determinants of behaviour change, including cognitive, affective, social and environmental influences (30). To ensure flexibility, the interview guide was updated during the data collection phase. Interviews were audio recorded and conducted by one or two interviewers (MC, DG, and/or RH) at the participant’s preferred location. Most interviews were conducted at the participants’ home, while a few were conducted at the public health service or a public place. No established relationship existed between the researchers and the participants before the interview, except for interactions during recruitment. The interviewers were a medical student (MC), a public health nurse with interview-experience (DG) and a health scientist (MSc) (RH). The interviewers were aware of potential personal bias and assumptions associated with their roles within a public health service, which ensured that these would not influence the data collection.

Data saturation was determined based on two criteria: ensuring sample diversity representative of the regional populations according to the previously mentioned criteria and reaching thematic saturation (32).

### Procedure

During recruitment, participants were informed face-to-face or by telephone on study details such as goals and confidentially of collected information, and the background of the researcher(s) by a researcher that attended the interviews. The interviews were carried out between May 2024 and February 2025 and lasted approximately one hour (ranging from 35-105 minutes). First, a test interview was conducted, recorded, and reviewed with MC, DG and a PhD candidate experienced in conducting qualitative interviews. Additionally, the first interview was reviewed by IM, who is experienced in qualitative research. Testing behaviour was assessed prior to the interviews and was further discussed during the interviews based on their responses. Level of education was defined according to the Central Statistical Office of the Netherlands (CBS) (33). After obtaining informed consent, the interviews started with introductory questions aimed at assessing the knowledge of avian influenza and testing, and their opinion on testing in a general sense. Subsequently, a short one-minute animation was shown to all participants illustrating a hypothetical AI outbreak. The animation started with a short introduction about avian influenza. After this, two specific stages simulated the early phase of a pandemic were described. This description covered 3 subjects: disease severity, level of uncertainty regarding the disease characteristics, and the desired behaviour of the population. Each participant was presented with the entire animation: the introduction and the two stages. In between the different parts of the animation and following the animation, participants were interviewed a subsequent part of the interview guide (see supplementary information). Memos were documented and updated during and after the interviews.

### Avian influenza animation

The animation started with an introduction of AI how it is commonly known: a bird flu that can occasionally infect humans, causing symptoms ranging from those comparable to a mild cold to more severe symptoms such as shortness of breath and pneumonia. Part one visualised the initiation of a nearby AI outbreak among poultry, followed by reports of individuals with symptoms of AI. The animation describes that human-to-human transmission has not yet been confirmed, but that sick individuals are advised to follow preventive measures and get tested. Part two reflects a subsequent phase in the outbreak, with increasing numbers of human infections and hospital admissions across all ages. Most of them did not have contact with poultry. In this phase, human-to-human transmission is confirmed, but a vaccine is not yet available. Recommendations of public health authorities include stricter adherence to preventive measures and testing. It should be noted that the animation refers to infections in the general population, without explicitly stating that the respondent will be affected. The overall purpose of the different steps is to imitate the development of a real emerging outbreak so that responses are not contingent on one specific phase.

### Data processing and analysis

The conducted interviews were transcribed verbatim. Transcripts were pseudonymized and recordings were directly deleted following transcription. Subsequently, a deductive thematic analysis approach was used to code the data, and themes were organized in the Capability Opportunity and Motivation model of Behaviour (COM-B). The COM-B model explains behaviour through three overarching components of behaviour: capability, whether one is able to perform the behaviour; motivation, whether one wants to do so; and opportunity, whether the environmental circumstances allow the behaviour (34, 35). Coding of all transcripts was conducted by one researcher (RH) using Atlas.ti version 8. To ensure consistency in coding and enhance coding credibility, a coding tree, was developed and systematically applied. 25% of interviews were independently double coded by a second researcher (DG). Transcripts were examined repeatedly to identify patterns and variations in the data (36). Identification of themes were both performed in advance and newly derived from the data. The codes were adapted and refined repeatedly as the researchers became more familiar with the data. In case of coding discrepancies, central issues or new emerging themes, a third researcher (LG) was involved. In addition, quotations of identified themes were discussed and checked by RH and LG (36).

## Results

### Sample characteristics

Seventeen participants were interviewed. These interviewees formed a heterogenous group in terms of age (median= 44; range 20-81), gender, education level, testing behaviour and country of birth of themselves and their parents (see Table 1 and Table S1 in the Appendix for sample characteristics and demographic information of the participants respectively).Twenty-two participants who had initially registered did not participate in the interviews because they withdrew or because a sufficient number of participants with similar characteristics was obtained.

**Tabel 1.** Sample characteristics reported by the participants (n = 17).

| Characteristic | Median<br>(range) or n<br>(%) |
| --- | --- |
| **Age, years** | 44 (20-81) |
| **Gender** |  |
| Female | 12 (71%) |
| Male | 5 (29%) |
| **Education level** |  |
| Lower | 7 (41%) |
| Middle | 4 (24%) |
| Higher | 6 (35%) |
| **Country of birth** |  |
| Netherlands | 10 (59%) |
| Other | 7 (41%) |
| - Suriname | 3 (43%) |
| - Turkey | 2 (29%) |
| - Iraq | 1 (14%) |
| - Germany | 1 (14%) |

Participants described several factors underlying their testing behaviour during a hypothetical outbreak with AI. We primarily report on these organised according to the COM-B model. However, need for autonomy and freedom of choice were identified as key themes, warranting separate discussion because of their significant and overarching role, and the fact that these are not captured within one component of the COM-B framework.

### Need for autonomy and freedom of choice

Participants with a lower willingness to test emphasized the importance of experiencing the need for autonomy or freedom to decide to test. They desired the possibility to make well-informed decisions (e.g., about when and how to test) and wanted to act independently within their own environment without being influenced by other factors such as requirements set by governmental authorities or social pressure. Having the freedom to choose would make testing more acceptable, whereas a decreased sense of autonomy would discourage testing: *“It is mainly the fact that people feel that things are required, and that, to me, feels like something being imposed on everyone. At that point, I no longer want to (get tested)” (R7).* Pressure from the government, such as restrictions aimed at enforcing testing, was especially met with criticism: *“Just stop it, all of you, I am not going to do that [..]. Look, I think every person is responsible for their own health, so it should never come from the government anyway.”* (R2). Participants stated that, rather than complying with externally imposed testing restrictions, they would adopt preventive measures, such as staying at home or social distancing: *“I’m not going to my mother, I’m just staying at home. […] When I start feeling better, I’ll go out again. But that whole testing we had, that constant preventive testing… […] I thought: I’ve been tested like crazy for my job. What was the point of that?” (R9).* In turn, this could be a reason to postpone testing or not test at all. Together, these results show the fundamental role of autonomy for willingness to test.

### Motivation

Different motivational reasons for getting tested were mentioned, such as: perceived usefulness of testing protection of own and others’ health, perceived sanctions, sanctions, trust, and psychological reassurance.

#### Disease severity and usefulness of testing

Participants’ attitudes and expectations regarding disease severity played a critical role in their decision to test. Participants indicated that their willingness to test depends on how severe they expect the disease to be or by how serious they perceived it at that time. Relatedly, the presence or absence of symptoms would influence testing behaviour. Participants reported that they would test when experiencing symptoms, either immediately or after postponing in case of very mild symptoms. Some participants with a lower willingness to test stated that they would only test when they perceived the symptoms as severe or unusual. Mild or familiar symptoms (i.e., comparable to a common flu, and referred to the symptoms from the animation) were not considered a reason to test: *“Yes […], if you are seriously ill and in the hospital, then think you would definitely want to know what is going on and where it is coming from […] and then I might want to know that myself too. But not if I am like, well, you know, I just feel a bit flu-ish, few days of rest and I will be fine. I think it also depends on how serious the symptoms are.”* (R12). Increased disease severity in the second part of the animation did not lead to new insights.

Another important factor was the perceived usefulness of testing, which was doubted by participants with a lower willingness to test. Some participants questioned the usefulness of testing because the lack of expected treatment options for themselves: *“There is no vaccine yet, there is no treatment yet. There is basically nothing at all. Sow what is the point for me, in the situation that I just saw, what do I actually gain form getting tested?”* (R13). Others questioned the usefulness because of a low perception of disease severity. Whereas participants mainly focused on individual-level considerations, population-level benefits, like pressure on hospital care, preventing lockdowns and their associated societal effects, were rarely mentioned.

#### Testing for own and others’ health

Mainly among participants with a higher willingness to test, the protection of the participants’ own health was mentioned as a reason to test: *“I would do it for myself, because […] if you have those symptoms, then you should just go get tested for your own sake, and for my wife too. Her health is really important to me, because she has a weakened immune system due to her diabetes.”* (R15). Protecting others in one’s social network emerged as the primary motivation for testing. Participants specifically emphasized the intention to test for people within their immediate circle, such as family or close friends: *“Look, if they are your soulmates, then I would say yes. But if it is just some casual acquaintance, oh come guys, who cares, do not do it”* (R2). Some participants indicated that they would also test to protect vulnerable people outside their circle, for example, elderly or hospitalized individuals.

Besides protective motives, participants indicated that testing would provide them psychological reassurance, such as a feeling of security or peace of mind when having a negative test result. This feeling of reassurance was sometimes said to be related to someone’s social network.

#### Perceived sanctions

Some participants experienced the prospect of quarantine or isolation, while waiting for the test result or after a positive test, as a sanction. To them, this formed a barrier for testing behaviour: *“Yeah, if I do not know, I would rather just keep it that way, and I will make sure myself that I do not go near vulnerable people. But if you know, then you are not allowed to go anywhere anymore.”* (R13).

#### Trust in governmental authorities and past experiences from the COVID-19 pandemic

Lower trust in the political government and governmental organisations such as the National Institute for Public Health and the Environment (RIVM) emerged as a recurring theme in participants with a lower willingness to test. Specifically, these participants expressed scepticism or lack of faith towards governmental authorities, influencing their testing behaviour. However, some participants described the Public Health Services in a more positive way than the government. This was reflected in their quotes illustrating their willingness to consider testing when approached by the Public Health Services: *“The government should just keep its mouth shut, to put it bluntly. [..] Then I would be much more willing to do it (testing at the Public Health Services). Yeah, sure, I would think about it a couple of times, but I would still do it sooner.”* (R2). In relation to lower trust in governmental authorities, some participants expressed a lack of clear information on testing, mainly based on the COVID-19-pandemic. Notably, participants often referred to experiences from the COVID-19-pandemic whilst discussing their testing behaviour during a hypothetical AI outbreak. This included reflections on trust, policy and preventive measures implemented during the COVID-19-pandemic, such as vaccination strategies and conditions regarding access to places.

### Opportunity

Multiple determinants within the opportunity domain were identified underlying testing behaviour, including external conditions imposed by the workplace or governmental authorities, external requests from governmental authorities and the perceptions of distance to the testing site and travel time.

#### External restrictions and requests

A common theme was conditions imposed by the workplace or governmental authorities, such as restrictions that have direct consequences, like being unable to go to work or attend an event without a negative test result. Only under exceptional circumstances, it was expressed that these conditions could increase testing behaviour, although this was often met with resistance: “*If my work says it is required, then I will do it [..] Then I still will not like it though [..], because I don’t think it is okay that people have to force me to do that.*”(R7). In line with this, participants indicated that external restrictions can create more resistance: *“I am not going to get tested three times a week just to be able to play sports […] So then we were not longer welcome […] Suddenly you are no longer part of society […] Very strange, but there is no one who is going to force me.”* (R9). In contrast, external requests from governmental authorities that leave more room for freedom of choice, such as invitations by letter or a clear testing advice in a newspaper, would encourage participants to test.

#### Distance to testing location

Distance and time to travel to the test site was related to testing behaviour: a shorter distance and/or travel time would facilitate participation, while a longer distance or travel time was perceived as a potential barrier. Some respondents reported being dependent on others for transportation to a test site.

### Capability

Within capability, one theme underlying testing behaviour was identified: knowledge.

#### Knowledge

The participants seemed to have varying degrees of knowledge regarding testing and AI. Notably, poor knowledge corresponded with a lower perception of disease severity and/or usefulness of testing. Specifically, participants had limited knowledge on seriousness of AI broader than individual or more immediate consequences. Additionally, the usefulness of testing besides for one’s own health (e.g. contact tracing, halting transmission) was not clear for all participants: *“Yeah, mostly because I think: if you’re not getting seriously ill, what is the point of testing to find out where the virus came from?”* (R11).

## Discussion

This qualitative study aimed to identify behavioural determinants of testing behaviour, particularly testing by a healthcare professional, based on a hypothetical human-to-human AI outbreak. Our results show that the decision to comply with testing during a hypothetical AI outbreak is a complex process with multiple behavioural factors involved. The most prominent identified themes were: need for autonomy and freedom of choice, external restrictions enforced by governmental authorities or employers, trust in governmental authorities, and past experiences from the COVID-19 pandemic. Participants mostly expressed matters of individual benefit and hardly included a public health perspective as an underlying reason for testing.

Our study demonstrates that a decreased sense of autonomy could hamper testing behaviour, while satisfaction of the need for autonomy could stimulate testing. Our findings align with prior research on other health behaviours, such as vaccination behaviour, indicating that decreased sense of autonomy reduces willingness to vaccinate, whereas satisfaction of the need for autonomy may have the opposite positive effect (37, 38). Consequently, the enforcement of external controlling factors directly undermines the experience of autonomy (39, 40). It is known that autonomy is negatively affected by enforcement of external controlling factors that do not reflect own values or interests (39, 40). Additionally, it has been shown that externally regulated behaviours tend to hinder the initiation of behaviour. This is problematic, as testing is a behaviour that likely encouraged or enforced externally and on top of that should likely be performed multiple times during a pandemic. Behaviours driven by autonomous motivation are known to persist longer (40–42).

Another important finding is the role of past experiences and perceptions that may have influenced current attitudes towards testing during a hypothetical avian influenza outbreak. This was demonstrated in our interviews, where participants frequently referred to COVID-19 experiences, such as vaccination and external restrictions set by governmental authorities. Experiences with past restrictions were sometimes said to encourage compliance to testing, but simultaneously frustrated the individuals’ sense of autonomy by being forced to test, which was also identified in previous pandemic preparedness studies. Correspondingly, other research suggested that engaging in pandemic preparedness provoked feelings of anger, resistance and opposition. These reactions were linked to a reduced freedom of choice experienced during the COVID-19 pandemic and discouraged people from taking action (43). Similarly, the role of past experiences was identified in the context of compliance to other preventive measures, such as vaccination and hygiene behaviours (44).

Trust in authorities emerged as a prominent recurring theme underlying testing behaviour, which is an important, but vulnerable and easily damaged, determinant in behaviour related to health risks, such as non-compliance to preventive measures. Notably, participants with a lower willingness to test often expressed scepticism or lack of faith towards governmental authorities when referring to testing behaviour. Low trust in authorities has been associated with low compliance to recommendations. This aligns with previous research showing that trust in authorities, including health agencies and governmental authorities, relates to willingness to adopt recommended preventive behaviours (44–47), including testing (19, 21). The central role of trust, combined with our observation that participants frequently referred to their experiences with COVID-19 to inform how they would respond to a novel outbreak, suggests that it is essential to find ways to promote testing that do not come at a cost of reduced trust.

Furthermore, our findings indicate that testing behaviour was primarily interpreted and discussed from an individual perspective rather than a public health perspective. During the interviews, participants framed testing decisions in terms of personal gain across most determinants, including disease severity, perceived usefulness of testing and psychological reassurance. The interview guide included a question about public health related motives for diagnostic testing in general but did not comprehensively assess it. Still, most participants did not mention possible public health reasons for testing like reducing pressure on hospital care, preventing lockdowns and their associated societal effects, or mitigating the economic impact of a pandemic. Previous evidence on this issue remains inconclusive as some previous research demonstrates that individuals prioritise self-benefit motives in preventive behaviour, especially when the perceived risk is high (48, 49), whereas others suggest that both self-benefit motives and social-benefit motives (50) or society-benefit solely (51) are strong predictors of compliance to preventive behaviour.

Finally, our study identified themes that previous studies have also identified in relation to COVID-19 testing behaviour. These themes include: 1) perceived severity of the disease (21–25, 44, 52), 2) perceived usefulness of testing (16, 18–22, 52), 3) accessibility of the test site (19, 26, 27), and 4) disease susceptibility of people within someone’s (close) social network and vulnerable people (16, 19, 21).

### Strengths and limitations

This study has several strengths. Firstly, we included a heterogeneous sample in our study, covering subpopulations in terms of age, educational level, and country of birth. The sample included a balanced distribution of varying degrees of willingness to test, providing valuable insights. Secondly, different measures were taken to facilitate openness and transparency of the participants to collect rich data. Trust was established trough personal and clear communication about the study aim, assurance of confidentially and explicit non-judgemental approach. In addition, interviews were conducted at the participants’ preferred time and location to facilitate comfort during the interview (53). Thirdly, during the interviews we used an animated video with clear B1 level information, which presented all phases of an early pandemic, enabling participants to better understand and engage in the hypothetical human-to-human avian influenza outbreak.

The main limitation of this study is the hypothetical approach. Participants’ reported or expected willingness to test may differ from their actual behaviour in real-life situations (18, 54), such as an actual human-to-human avian influenza outbreak. However, to prepare for future outbreaks we are by definition restricted to hypothetical scenarios. Moreover, the value of hypothetical scenarios has been demonstrated in other research (55). A second limitation is that qualitative interviews do not lend themselves well for gaining insights into the automatic processes like habits or impulses triggering decisions to test (56). Although we used a video animation and specific questions to probe automatic responses (e.g., questions about emotions), people are known to overestimate the role of deliberate processes on their decisions resulting in an likely underestimation of automatic responses (57). As it is likely that automatic processes are important for testing behaviour as well(58), different methods are required to capture this (56). The third limitation is that the study’s interviews were conducted by Public Health Service professionals instead of an external organisation. The perceived positive attitude of Public Health Service professionals towards testing may have influenced the conversation, particularly in participants with a lower willingness to test. Similarly, it cannot be excluded that participants were more positive about their trust in the Public Health Service because of social desirability. However, we noticed that participants were open and transparent in sharing their experiences, views and opinions, appeared unhampered in the rough language they used, and seemed comfortable in their chosen interview setting.

### Implications

We propose a number of key considerations when designing testing strategies. First, it is important to build and maintain trust, even in crisis situations when decision-making requires balancing between uncertainty and urgency demanding rapid action. Trust can be impacted by different determinants including lack of (access to) clear information (59, 60). Previous studies showed that a lack of clear information on testing negatively affected the willingness to engage in testing (16, 27). In our study, lack of clear information was mentioned in the context of trust and not particularly in the context of testing. Research has shown that when policy making is perceived as transparent, people are more likely to trust government decisions (59) and follow risk communication advise (61). Thus, transparency in communication should be prioritized to allow insights in governmental decisions (59, 62). Moreover, different sources may be trusted differently (63). Our study for example revealed that the Public Health Services were not always perceived as part of the governmental authorities and were sometimes granted a higher level of trust. Altogether, key elements such as building trust, considering who serves as the information provider, and ensuring transparency of information need to be taken into account (47, 59).

Second, autonomy is a crucial factor in designing testing strategies. Although autonomy is known to be undermined by the imposition of external controlling factors (39, 40), from a biomedical perspective restrictive measures may still be warranted in outbreak situations to protect public health. In such cases, a key issue is how communication can acknowledge individual perspectives and decision making capacities (47). Offering the opportunity to choose could be part of the strategy, such as allowing alternative preventive measures. In addition, ensuring easy access to transparent and relevant information is essential to enable individuals to make their own informed decisions (16, 27, 47). These insights can guide future research on autonomy-supportive communication.

Finally, it is necessary to reflect on how motives to stimulate testing are communicated, as our study revealed that participants are most aware of possible individual gains of testing rather than public health benefits. One way could be to leverage the already existing individual motives as they seem to play a prominent role in shaping testing behaviour. Conversely, an alternative approach could be to increase awareness of the public health benefits as our study results show that there is room for improvement on this perspective. However, it is then still not clear whether this awareness would translate to increased willingness to get tested. As previous evidence seems inconclusive at this point, more research is needed to determine how self-benefit and societal-benefit should be included in communication (48–51).

## Conclusions

This study demonstrates that testing behaviour during a hypothetical human-to-human AI outbreak is a complex process shaped by multiple factors, such as autonomy and external restrictions set by governmental authorities or employers, or prior experiences with testing. The results suggest that determinants of testing behaviour are not disease-specific, but may be more universally applicable. The insights of this study can inform the design of communication and testing strategies that are tailored to the populations’ motivations and preferences. This is necessary because solely focussing on containing the outbreak through among others external restrictions, can directly and negatively impact trust and experienced autonomy. In return, these experiences can directly impact behaviour during a future infectious disease outbreak. Hence, future research is needed to determine how these findings can be translated into effective communication and how trust in authorities can be built. Key considerations for informing communication strategies include balancing need for autonomy with external restrictions, rebuilding trust in institutions, providing transparency in information and policy processes.

## Data Availability

Due to the sensitive nature of the qualitative data and the potential for identification, we manage data sharing carefully. Any request for access to individual data are assessed by the research team.

## Abbreviations

AI: Avian Influenza
CBS: Central Statistical Office of the Netherlands
COM-B: Capability, Opportunity and Motivation model of Behaviour
METC LDD: The Medical Research Ethics Committee Leiden and The Hague
RIVM: National Institute for Public Health and the Environment
TDF: Theoretical Domains Framework.

## Acknowledgements

We would like to thank Mia Curtis (MC), who at the time of the study was medical student at Leiden University, for conducting several interviews for this study. We also thank Rukiye Turkeli, PhD candidate at the Department of Public Health and Primary Care Campus The Hague, Leiden University Medical Centre, for conducting the pilot interview with us. Finally, we are grateful to all participants for their time and valuable contributions to this study.

## Declarations

### Author contributions

AS, RH, LG, HB, MP, IM, LV, DG, CK and MM contributed to the conception and design of the study. Data were collected by DG and RH, and the coding process was performed by RH, DG and LG. Data analysis was conducted by RH and LG, with additional interpretation provided by AS, HB, IM, MP, CK, and MA. This research was supervised by AS, LG and MA. The original draft of the manuscript was prepared by RH, LG, AS and MA. All authors contributed to the revision of the manuscript and approved the final version.

### Ethical considerations and consent to participate

The Medical Research Ethics Committee Leiden and The Hague (METC LDD) determined that this study does not meet the criteria of the Medical Research Involving Human Subjects Act and is therefore classified as non-WMO research (N24.031). All participants provided informed consent prior to participation in the interviews. Confidentiality and pseudonymity were ensured according to consultations with a privacy officer of the Municipality of The Hague.

### Consent for publication

All participants provided consent for the publication of pseudonymized data.

